# Does the application of Robotics improve outcomes of pedicle screw insertion in spine surgery compared to conventional fluoroscopy guidance? A protocol for Systematic Review and Meta-Analysis

**DOI:** 10.1101/2021.12.08.21267465

**Authors:** Vishal Kumar, Vishnu Baburaj, Prasoon Kumar, Sarvdeep Singh Dhatt

**Author notes:** <u>*Correspondence to:*</u> Dr. Vishnu Baburaj, Senior Resident, Department of Orthopaedics, Postgraduate Institute of Medical Education and Research, Chandigarh, India. 160012.

## Abstract

**Background:** Pedicle screw insertion is routinely carried out in spine surgery that has traditionally been performed under fluoroscopy guidance. Robotic guidance has recently gained popularity in order to improve the accuracy of screw placement. However, it is unclear whether the use of robotics alters the accuracy of screw placement or clinical outcomes.

**Objectives:** This systematic review aims to compare the results of pedicle screws inserted under fluoroscopy guidance, with those inserted under robotic guidance, in terms of both short-term radiographic outcomes, as well as long-term clinical outcomes.

**Methods:** This systematic review will be conducted according to the PRISMA guidelines. A literature search will be conducted on the electronic databases of PubMed, Embase, Scopus, and Ovid with a pre-determined search strategy. A manual bibliography search of included studies will also be done. Original articles in English that directly compare pedicle screw insertion under robotic guidance to those inserted under fluoroscopy guidance will be included. Data on outcomes will be extracted from included studies and analysis carried out with the help of appropriate software.

## 1. Background

Pedicle screw insertion is routinely carried out in spine surgery for several indications including traumatic, infective, degenerative and neoplastic conditions and spinal deformities. Conventionally, it has been carried out with free-hand techniques under fluoroscopy guidance where anatomical landmarks are used to guide pedicle entry site and direction. However, the use of this technique has frequently been associated with misplaced screws [1]. Adjacent neurovascular structures and visceral organs are prone to injury by such malpositioned screws. Technical advancements have led to the application of navigation and robotics in spine surgery to improve the accuracy of screw placement.

## 2. Need for review

Whether the use of robotic guidance leads to improved accuracy and clinical outcomes remains controversial. Some studies report improved screw accuracy, reduced rate of complications and intra-operative radiation exposure with robotic guidance [2-7]. Some other studies, including two RCTs and a meta-analysis have concluded that Robotics have no added benefit over free-hand placement under fluoroscopy guidance [8-11].

### Objective

To compare the outcomes of pedicle screws inserted under fluoroscopy guidance with those inserted under robotic guidance.

## 3. PICO framework for the study

a. Participants : Adult human subjects undergoing Spine surgery
b. Intervention : Pedicle screws inserted under robotic guidance
c. Control : Pedicle screws inserted under fluoroscopy guidance
d. Outcome : Accuracy of screw placement, mean surgical duration, blood loss, radiation exposure, complication and screw revision rates

## 4. Methods

This systematic review and meta-analysis will be done according to the Preferred Reporting Items for Systematic Reviews and Meta-analysis (PRISMA) guidelines.

a. *Review Protocol* A protocol of the review will be prepared as per the PRISMA-P guidelines.
b. *Eligibility Criteria* Original research on human adult subjects having spinal pathology undergoing spine surgery with pedicle screw fixation under robotic guidance will be included. The studies should have a comparison group in which pedicle screws are inserted under fluoroscopy guidance. The articles must report on the clinical and radiological outcome parameters. Studies in languages other than English, lower evidence studies such as case reports, case series, animal, cadaveric and biomechanical studies will be excluded.
c. *Information Sources and Literature search* Electronic databases of PubMed, Embase, Scopus, and Ovid will be searched using the keywords “Robot* AND (Spine* OR (Pedicle Screw))” for studies in English published from inception to date of search. A bibliography search of included studies will also be carried out for more potentially eligible articles.
d. *Study Selection* Two authors will separately go through the title and abstract of the search results to narrow down studies using the inclusion and exclusion criteria. In case of any doubt, the full text of the study will be obtained and an assessment made after discussion with all the authors.
e. *Data Collection and Data Items* Data from eligible studies will be extracted on excel spreadsheets, which will be cross-checked for accuracy. The following data will be collected:
  - Name of first author and publication year
  - Study design
  - Type of robot used
  - Number of participants and their demographic data
  - Mean operating time
  - Accuracy of screw placement
  - Blood loss
  - Radiation exposure
  - Rate of complications and screw revision.
f. *Outcome measures* The outcome measures that would be considered for analysis are as follows:
  - Mean operating time
  - Accuracy of screw placement
  - Blood loss
  - Radiation exposure
  - Rate of complications
  - Rate of screw revision.
g. *Data Analysis and Synthesis* Both qualitative and quantitative synthesis will be performed, if adequate data is obtained. Meta-analysis would be conducted to compare the pooled estimate of outcomes between robot guided and fluoroscopy guided techniques if reported by more than three studies. RevMan version 5.4 (computer program) will be used for analysis. A fixed or random-effects model will be chosen based on the amount of heterogeneity. 95% confidence intervals will be used, and results would be depicted using forest plots.
h. *Assessment of Risk of Bias* The methodological index for non-randomized studies (MINORS) tool will be used to assess bias in observational studies, and Cochrane risk of bias tool will be utilized for randomized controlled trials [12, 13].

## Data Availability

All data produced in the present work are contained in the manuscript

